# Temporal features of the built environment and associations with drowning mortality: A global satellite-based analysis

**DOI:** 10.64898/2026.04.19.26351237

**Authors:** Ryan Essex, Samsung Lim, Jagnoor Jagnoor

## Abstract

**Background:** Drowning remains a major global public health challenge. This study examined whether the timing and trajectories of urbanisation—beyond the current built environment—are associated with subnational drowning mortality.

**Methods:** We linked satellite-derived measures of built-environment change (GHSL), population crowding (WorldPop), surface water exposure (JRC Global Surface Water), and infrastructure proxies (VIIRS/DMSP nighttime lights) to GBD 2021 drowning mortality estimates across 203 ADM1 regions in 12 countries (2006–2021; 3,248 region–year observations). Temporal predictors captured recent expansion, development “newness” (≤10-year built share), acceleration/volatility, and a crowding×growth interaction. We screened predictors using LASSO (10-fold cross-validation) and fitted mixed-effects models with region random intercepts. Distributed-lag models tested temporal precedence and development age, and income-stratified models assessed heterogeneity.

**Results:** Adding temporal predictors improved fit beyond contemporaneous built-environment measures (ΔAIC=177; ΔBIC=147). In adjusted models, crowding×growth was strongly positively associated with drowning mortality, and a higher share of recent development was associated with higher mortality. Lag models showed a development age gradient: older built environment was most protective. Associations differed by income group, with several key coefficients reversing sign across strata.

**Discussion:** Drowning mortality appears shaped by development histories as well as present-day conditions, with risk concentrated in rapidly changing, dense settings and the newest built environments. Cross-context heterogeneity suggests mechanisms and prevention priorities are unlikely to be uniform.

**Conclusions:** Development timing and trajectories help explain subnational drowning mortality beyond current built form alone. Prevention and planning should prioritise transition-period safety strategies in newly developing and rapidly densifying areas.

## Background

Drowning remains a major global public health challenge, responsible for millions of deaths over the last decade (World Health Organization, 2024). While drowning risk factors have traditionally focused on individual behaviours and supervision, the built environment represents a critical but understudied determinant of population-level drowning mortality, particularly in contexts undergoing rapid urban transformation. The literature that exists in this space points to the fact that the built environment matters, with rural areas consistently showing higher drowning rates than urban areas in studies from Canada [1], the US [2], and China [3]. However, this rural-urban divide hardly captures the dynamic processes of urbanisation and development. Rapid unplanned urbanisation, for example, may create or promote hazards through informal waterfront settlements, inadequate drainage and flooding infrastructure, and unsafe water access points. The United Nations projects that 68% of the world’s population will live in urban areas by 2050, with 90% of this growth occurring in Asia and Africa [4]. This means that many settings will experience not just higher levels of urbanisation, but rapid transitions in their built environment, shifts that may carry distinct drowning risks that differ from those in stable urban or rural contexts.

Despite the importance of these dynamics, the literature has focused almost exclusively on cross-sectional comparisons, with very little attention to how the pace and timing of urbanisation influence drowning risk. We know little about whether recently developed areas carry different risks than established urban settlements, or whether there are lag periods during which protective infrastructure effects emerge. These gaps are particularly pressing given evidence from other fields that infrastructure benefits often take time to materialise [5]. In injury prevention more broadly, structural interventions are recognised as critical [6], but the temporal dynamics of how these interventions work, and whether newly built environments experience transition periods of elevated risk before safety systems mature, remain largely unexplored in the drowning prevention literature. Understanding these temporal processes has direct implications for prevention.

Satellite-derived data now enables examination of built environment changes over time. Global settlement data provides measures of urban expansion, infrastructure development, and population redistribution at fine spatial resolutions and across multiple time points [7]. Combined with water exposure data [8] and longitudinal mortality estimates, this data allows the testing of whether past urbanisation predicts current drowning rates, and whether protective effects emerge gradually as infrastructure matures. In this study we examine temporal dynamics of urbanisation and drowning mortality, using satellite-derived measures linked to subnational estimates from the Global Burden of Disease study.

## Research Aims

Building on previous cross-sectional analysis of contemporary built environment characteristics [9], this study investigates whether urban growth patterns over time, historical development patterns, and the timing of infrastructure provision influence drowning risk beyond the current built environment state alone. Specifically, we sought to: (1) quantify temporal change effects by testing whether urbanisation pace, recent development, and crowding-growth interactions predict drowning mortality independently of current built environment characteristics; (2) test temporal precedence (whether earlier urban development is linked to later drowning rates) through distributed lag models examining whether historical urbanisation levels and development predict current drowning risk; (3) assess infrastructure timing mechanisms by testing whether rapid development outpacing service provision elevates drowning risk; and (4) examine heterogeneity in temporal dynamics by testing whether effects differ across country income levels.

## Methods

### Data sources and extraction

Built environment data were extracted from Global Human Settlement Layer (GHSL) [7], Visible Infrared Imaging Radiometer Suite and Defense Meteorological Satellite Program (VIIRS/DMSP) nighttime lights [10, 11], WorldPop [12] population estimates, and Joint Research Centre (of the European Commission; JRC) Global Surface Water [8] datasets using Google Earth Engine (GEE) [13]. GHSL Built-Up Surface provided 100 m built-area estimates at 5-year intervals (1975–2025). For each study year, we extracted built area for the nearest epoch and for 5 and 10 years prior (e.g., 2020, 2015, 2010), enabling calculation of growth, change magnitude, and lagged development measures. WorldPop provided annual 100 m population estimates, used to derive population crowding within built-up areas. Nighttime lights were drawn from DMSP-Operational Linescan System (OLS) (2006–2013) and VIIRS Day/Night Band (2014–2021) as proxies for infrastructure and service provision. Water bodies were derived from JRC Global Surface Water (1984–2021) to identify built areas near surface water. For computational efficiency, ADM1 geometries were simplified to a 5 km tolerance and buffered by 50 km during extraction to reduce edge effects in distance and aggregation calculations. Analysis focused on subnational regions (ADM1 level) with drowning mortality estimates (age standardised rate, which includes unintentional drowning events) from the Global Burden of Disease (GBD) 2021 study [14] that matched Global Administrative Unit Layer (GAUL) 2015 [15] administrative boundaries (n= 203 regions across 12 countries covering 2006-2021). Regions were therefore included where drowning data was available and where boundaries had stayed relatively stable over the study period. Countries included in this analysis were Brazil, Ethiopia, India, Iran, Italy, Japan, Kenya, Mexico, Norway, Pakistan, South Africa, United States of America.

### Variable construction

A summary of all variables derived in this study is included in Table 1 and a brief description of each is provided below.

**Table 1.** Summary and description of variables included in modelling.

| Variable | Models used | Construction formula | Interpretation (expanded) |
| --- | --- | --- | --- |
| **Built change ratio (5-year proportional growth)** | LASSO; Main model | $(\text{Current built area} - \text{Built area 5 years ago}) \div (\text{Built area 5 years ago})$ | **Scale-free measure of recent urban expansion intensity.** Higher values indicate faster proportional growth of built-up area over the preceding 5 years; values near zero indicate little recent expansion; negative values (if present) would indicate contraction or reclassification. Captures *how much* the built footprint has expanded recently relative to its baseline size. |
| **Growth acceleration (change in growth rate)** | LASSO; Main model | $[(\text{Current built area} - \text{Built area 5 years ago}) \div 5] - [(\text{Current built area} - \text{Built area 10 years ago}) \div 10]$ | **Whether urbanisation is speeding up or slowing down.** Positive values indicate acceleration (recent annualised growth exceeds longer-term annualised growth); negative values indicate deceleration (growth is slowing). Separates *trend shifts* from overall growth level. |
| **Growth volatility (instability in growth dynamics)** | LASSO; Main model | Absolute value of $\{[(\text{Current built area} - \text{Built area 5 years ago}) \div 5] - [(\text{Current built area} - \text{Built area 10 years ago}) \div 10]\}$ | **Magnitude of fluctuation in urban growth, regardless of direction.** Higher values indicate more “boom–bust” dynamics or instability (rapid changes in growth rate); lower values indicate steadier, more predictable development trajectories. Treats sharp acceleration and sharp deceleration as similarly “volatile.” |
| **Recent development ratio ($\leq 10$-year built share)** | LASSO; Main model; Lag Model 3 | $(\text{Current built area} - \text{Built area 10 years ago}) \div (\text{Current built area})$ | **How “new” the current urban fabric is.** Values closer to 1 indicate a larger proportion of the present-day built environment was added within $\sim 10$ years (newer development); values closer to 0 indicate a predominantly older built environment (more established urban form). Conceptually captures the *recency* of development rather than its rate alone. |
| **Crowding × growth (interaction: density-sensitive expansion)** | LASSO; Main model | Natural logarithm of $(\text{Crowding}) \times [(\text{Current built area} - \text{Built area 5 years ago}) \div (\text{Built area 5 years ago})]$ | **Tests whether rapid expansion is more consequential in already dense settings.** Higher values reflect **fast growth occurring where crowding is high**, consistent with compressed development pressure; lower values reflect slower growth and/or less crowded environments. Operationalises the hypothesis that growth may carry different implications depending on baseline density. |
| **Historical urbanisation (t–10 urban extent)** | Lag Model 1 | $(\text{Built area 10 years ago}) \div (\text{Total land area})$ | **Baseline urban extent a decade earlier.** Higher values indicate regions that were already more built-up at t–10 (more established urbanisation); lower values indicate less built-up regions at baseline. Used to distinguish *level effects* (existing urbanisation) from *change effects* (recent growth). |
| **Recent urbanisation change (5-year change in urban extent)** | Lag Model 1 | $(\text{Current \% built area}) - (\% \text{ built area 5 years ago})$ | **Absolute change in built-up share over the prior 5 years.** Positive values indicate increasing urban extent (expansion); values near zero indicate stability. Complements the proportional “built change ratio” by expressing change in percentage-point terms (more interpretable as absolute change in land share). |
| **Old built environment (>10-year built area)** | Lag Model 2 | Built area 10 years ago | **Amount of mature built environment.** Higher values indicate a larger established built footprint (development older than ~10 years), typically assumed to coincide with more settled land-use patterns and, plausibly, more mature infrastructure and risk governance. Anchors the “age structure” decomposition of built environment. |
| **Mid-age additions (5–10-year additions)** | Lag Model 2 | $(\text{Built area 5 years ago} - \text{Built area 10 years ago})$ | **Built area added 5–10 years ago.** Captures development that is neither very recent nor long-established—potentially a transition period where infrastructure and safety systems may be partially adapted. Helps test whether risk associations differ by age of development. |
| **Recent additions (<5-year additions)** | Lag Model 2 | $(\text{Current built area} - \text{Built area 5 years ago})$ | **Built area added in the last 5 years.** Represents the newest development, where infrastructure, services, and safety governance may be least “caught up.” High values indicate substantial very recent expansion. |
| **Infrastructure lag in new development (nighttime-lights deficit per built area)** | Lag Model 3 | $(\text{Expected nighttime light per unit built area} - \text{Observed nighttime light per unit built area})$ | **A proxy measure for service provision not keeping pace with development.** Higher values indicate **newer / expanding built areas with relatively low nighttime light intensity per unit built area**, consistent with weaker infrastructure, services, or electrification relative to the built footprint; lower values indicate development occurring alongside comparatively stronger service provision. |
| **Recent development × infrastructure lag (interaction)** | Lag Model 3 | $[(\text{Current built area} - \text{Built area 10 years ago}) \div (\text{Current built area})] \times (\text{Infrastructure lag measure})$ | **Tests compounding risk where “newness” coincides with service deficits.** High values indicate places where a large share of the current built environment is recent **and** infrastructure lag is pronounced—consistent with rapid development without commensurate services. Interprets whether the association of new development depends on infrastructure conditions. |

### Temporal change variables

From the above data sources we derived five predictors capturing urbanisation dynamics: (1) Growth acceleration compared recent (5-year) growth with longer-term (10-year annualised) growth to indicate whether urbanisation was speeding up or slowing; (2) built change ratio captured proportional expansion over the prior 5 years; (3) recent development ratio measured how much of the current built environment was added in the last ∼10 years, distinguishing newer from long-established urban fabric; (4) growth volatility captured instability in development by comparing short– and longer-term growth rates; (5) finally, we tested whether rapid growth in already dense settings amplified risk using a crowding × growth interaction; crowding was log-transformed to reduce right skew.

### Lag structure variables

To assess whether timing mattered, we constructed lagged predictors (earlier urban conditions used to predict later drowning rates). Historical urbanisation (built-up share of land 10 years earlier) represented baseline urban extent 10 years prior, while recent urbanisation change (percentage change in built-up share over the last 5 years) captured the change in urban extent over the previous 5 years. We also divided built environment into old (>10 years), mid (5–10 years), and recent (<5 years) additions to test whether development age carried different drowning risk. These variables were derived to align with GHSL’s 5-year epoch structure (2000, 2005, 2010, 2015, 2020)

### Infrastructure timing variables

We examined whether service provision kept pace with development. Infrastructure lag captured contexts where recent expansion coincided with relatively poor infrastructure, proxied using nighttime light intensity per built area. A complementary interaction term (recent development × infrastructure lag) tested whether rapid development combined with infrastructure deficits compounded risk.

All temporal variables were standardised to z-scores (mean 0, SD 1) before analysis to enable effect-size comparability (so estimates are comparable across variables); scaling was performed on the full dataset prior to income-stratified models. All variables are summarised in Table 1, and further details on extraction and construction are reported in our previous study (9).

### Analytic Strategy

#### Overview

Analysis proceeded in five stages: (1) extraction and calculation of built environment variables using GEE; (2) variable selection through univariate association testing and LASSO; (3) main model incorporating temporal variables alongside contemporary predictors from our earlier study; (4) three lag-structured models to test specific hypotheses about temporal precedence and development timing; (5) income-stratified analyses to test whether temporal dynamics differ by development context. All analyses were conducted in R version 4.5.0 [16]. A complete list of R packages used is included in Table S1 (supplementary material).

#### Variable Selection

Least Absolute Shrinkage and Selection Operator (LASSO) regularisation was used to test whether temporal urbanisation measures added explanatory power beyond the contemporary built-environment predictors identified in our previous cross-sectional analysis. The LASSO model included (1) the eight contemporaneous built-environment variables from the prior study (see Supplementary Table S2) and (2) five temporal variables (growth acceleration, built change ratio, recent development ratio, growth volatility, and the crowding × growth interaction). We applied 10-fold cross-validation using the lambda.min criterion to identify temporal variables that improved fit. Following this screening step, we retained all variables for subsequent mixed-effects modelling (see results below).

#### Main Model

We fitted a mixed-effects linear regression with random intercepts for region (a region-specific baseline level to account for repeated measures) to assess whether temporal factors added explanatory power for drowning risk:

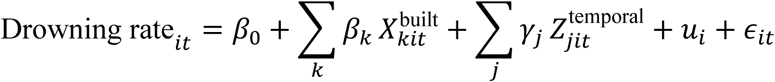

where *i* indexed region and *t* year. 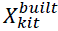 were contemporaneous built-environment predictors and 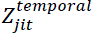 were temporal predictors. β_0_ was the overall intercept; β_*k*_ and γ_*j*_ were fixed-effect coefficients. *u*_*i*_ was a region-level random intercept (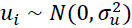) and ϵ_*it*_was the residual error term (ϵ_*it*_ ∼ *N*(0, σ^2^)).We compared this temporal-augmented model to the contemporaneous-only model (eight predictors; Supplementary Table S2) using AIC and BIC. Models were estimated using restricted maximum likelihood (REML).

#### Lag Models

We fitted three lag-structured mixed-effects models to test whether past urbanisation predicted current drowning risk and whether the timing of development mattered. All lag models included the same contemporary controls, log-transformed crowding and built area near water, to isolate temporal effects while accounting for established present-day risk factors. All models included random intercepts for region and were estimated using REML. In all lag specifications, *i*indexes region and *t*year; *t* − 10and *t* − 5denote 10-year and 5-year lags, respectively. Expressions such as *t* − 5 → *t*represent cumulative change over that interval. All βterms represent fixed-effect coefficients.

Model 1: Level vs change

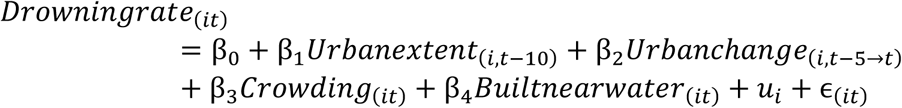

Model 2: Built environment by development period

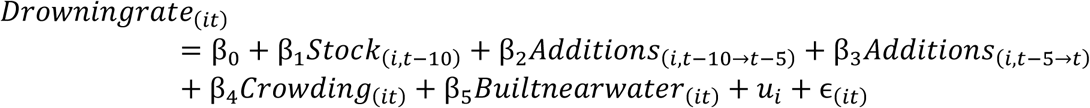

Model 3: Infrastructure timing

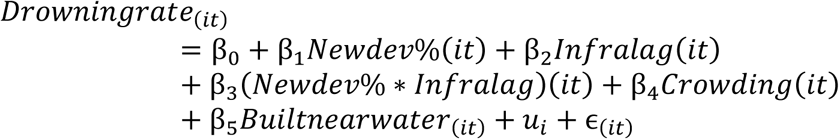

#### Income-Stratified Analysis

We fitted income-stratified versions of the main temporal model and lag Model 1 using 2021 World Bank income groups to assess whether temporal dynamics differed by development context. Separate mixed-effects models were fitted within each income group using observations from countries in that category, retaining region-level random intercepts. We did not estimate formal cross-stratum interaction terms due to limited sample sizes in some strata; instead, we present stratum-specific coefficients and confidence intervals to assess heterogeneity.

#### Model Diagnostics

We evaluated: (1) improvement from temporal predictors using AIC/BIC; (2) multicollinearity using variance inflation factors (VIF; >5 moderate, >10 severe); (3) residual diagnostics (Q–Q plots; residuals vs fitted; residuals vs predictors); (4) sensitivity to influential observations using Cook’s distance (cut-off 4/n) and refitting models; and (5) alternative specifications where feasible (e.g., random slopes for key predictors; alternative functional forms such as quadratic terms). We used two-tailed α = 0.05, emphasising effect sizes and confidence intervals. To test for residual spatial dependence beyond region-level random intercepts, we computed global Moran’s I on country-level mean residuals (N = 12) using k-nearest neighbour weights (k = 3, 5, 8) with analytical and Monte Carlo permutation tests (999 simulations), supplemented by Local Indicators of Spatial Association (LISA).

### Ethical approval

As this study utilised public data, ethical approval was not required or sought.

### Patient and public involvement

No patient or public involvement.

## Results

### Descriptive Statistics

The analysis included 3,248 region–year observations from 203 ADM1 regions in 12 countries over 2006–2021. Temporal urbanisation measures varied widely: the 5-year built change ratio averaged 0.09 (SD 0.08; range 0.01–0.85) and the recent development ratio averaged 0.16 (SD 0.08; range 0.02–0.64). Growth acceleration was negative on average (−0.72 pp; SD 1.51; range −11.73 to 8.76), suggesting overall deceleration, while growth volatility averaged 1.09 (SD 1.27; range 0.00–11.73). The crowding × growth term was strongly right skewed (mean 83.32; SD 70.14; max 781.89). All descriptive statistics are included in Tables S3-5 and Figures S1 (supplementary material).

### Variable Selection

LASSO regularisation with 10-fold cross-validation retained all five temporal variables alongside the eight contemporaneous built-environment predictors from our earlier study (Table S6). In univariate models, four temporal variables were positively associated with drowning rates (all *p* < 0.001), with the strongest relationships observed for crowding × growth (rapid growth in already crowded areas) (β = 0.524; R² = 0.079), recent development ratio (share built within ∼10 years) (β = 0.517; R² = 0.077), and 5-year built change ratio (5-year proportional growth) (β = 0.496; R² = 0.070). Growth acceleration (recent per-year growth minus longer-term per-year growth) showed a smaller association (β = 0.250; R² = 0.018), while growth volatility (magnitude of change in growth rate) was not associated (*p* = 0.34; R² ≈ 0). LASSO coefficients differed from univariate estimates, consistent with shared variance and suppression among correlated predictors and contemporary built-environment controls. Notably, crowding × growth switched sign in the multivariable LASSO (LASSO β = −2.396), whereas 5-year built change ratio remained positive and increased in magnitude (LASSO β = 1.518). The fact that all five temporal variables were retained suggests they captured distinct components of urbanisation dynamics (directional change vs instability; proportional expansion vs the “newness” of current built environment), despite moderate conceptual overlap (Table S6).

### Main model

Adding temporal urbanisation variables to the contemporary built-environment model substantially improved fit (Table S6). The temporal-augmented model (13 predictors) reduced AIC from 4,260.50 to 4,083.46 (ΔAIC = 177.04) and BIC from 4,327.44 to 4,180.83 (ΔBIC = 146.61), indicating a large improvement despite the penalty for additional parameters. In the fully adjusted model, the strongest temporal effect was the crowding × growth interaction (rapid growth in already crowded areas) (β = 1.773, SE = 0.245, z = 7.25, p < 0.001), suggesting that rapid growth in already-crowded regions was associated with higher drowning rates. The 5-year built change ratio (5-year proportional growth) was strongly negative (β = −1.662, SE = 0.239, z = −6.96, p < 0.001), contrasting with its positive univariate association and consistent with suppression/shared variance with other temporal and infrastructure-related measures. Recent development ratio (share built within ∼10 years) was positive (β = 0.328, SE = 0.050, z = 6.58, p < 0.001), implying higher risk where a larger share of the built environment was newly developed. Growth acceleration (recent per-year growth minus longer-term per-year growth) was negative (β = −0.076, SE = 0.015, z = −4.98, p < 0.001) and growth volatility (magnitude of change in growth rate) showed a small negative association (β = −0.025, SE = 0.011, z = −2.20, p = 0.028). Among contemporary predictors, log crowding remained strongly negative (β = −1.490, SE = 0.085, z = −17.47, p < 0.001). Several other built-environment measures were also negative (built area near water, infrastructure lag, urban extent, development pattern, and nightlight CV), while the crowd–water interaction was positive (β = 0.325, SE = 0.060, z = 5.44, p < 0.001). Nightlight per built area was positive but not conventionally significant (β = 0.021, SE = 0.011, z = 1.88, p = 0.060). (Figure 1 and Tables S6).

**Figure 1.**
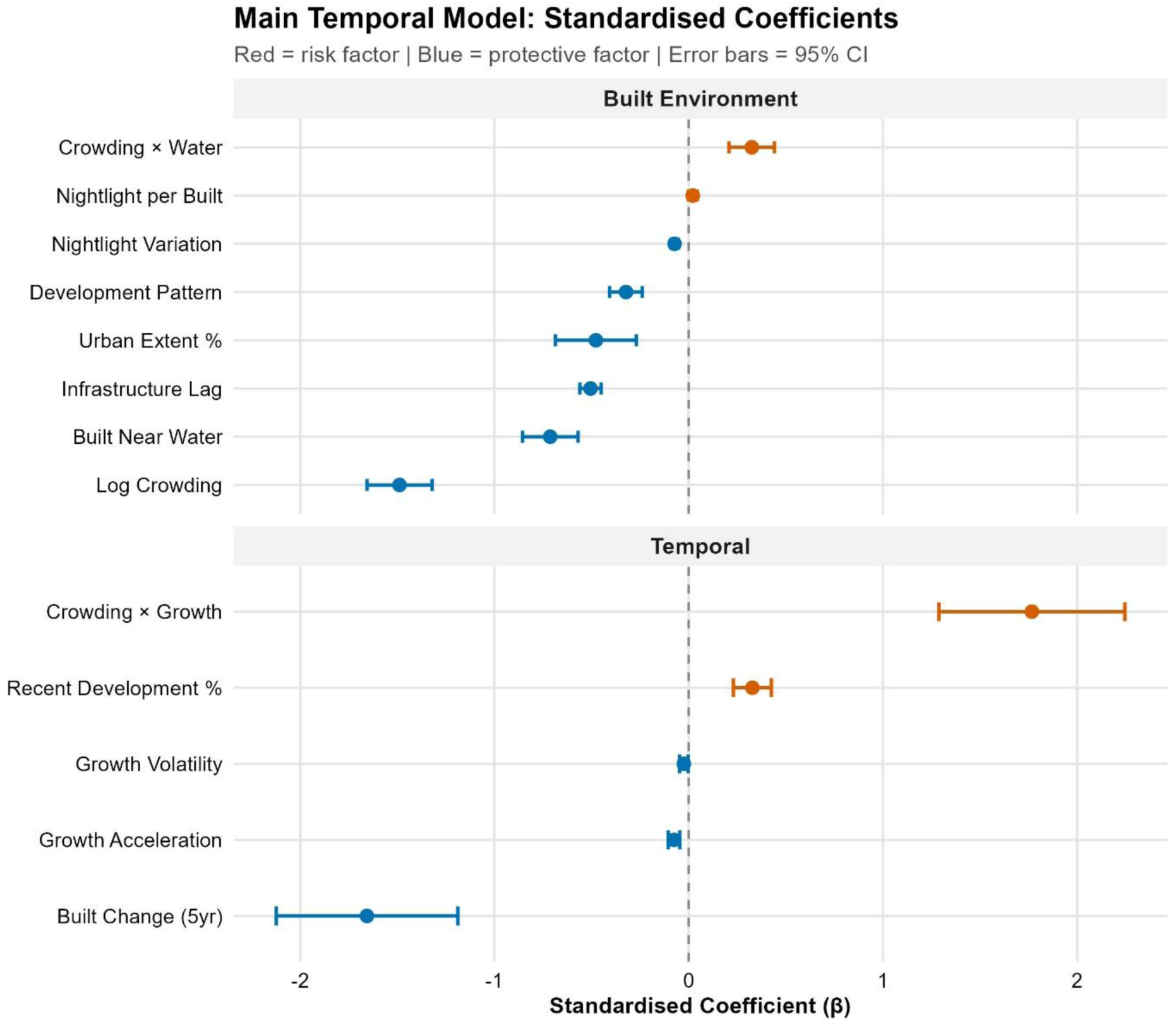
Main model results.

### Lag models

#### Level versus change effects

In the level-versus-change lag model, both historical urbanisation and recent change were negatively associated with drowning mortality (Table S7). Historical urban extent 10 years prior showed a large negative association (β = −1.538, p < 0.001), while recent 5-year urbanisation change was also negative but smaller in magnitude (β = −0.357, p < 0.001). This pattern suggests that longer-established urbanisation is more strongly associated with lower drowning risk than short-term changes, consistent with adaptation and the accumulation of protective infrastructure over time (Figure 2).

**Figure 2.**
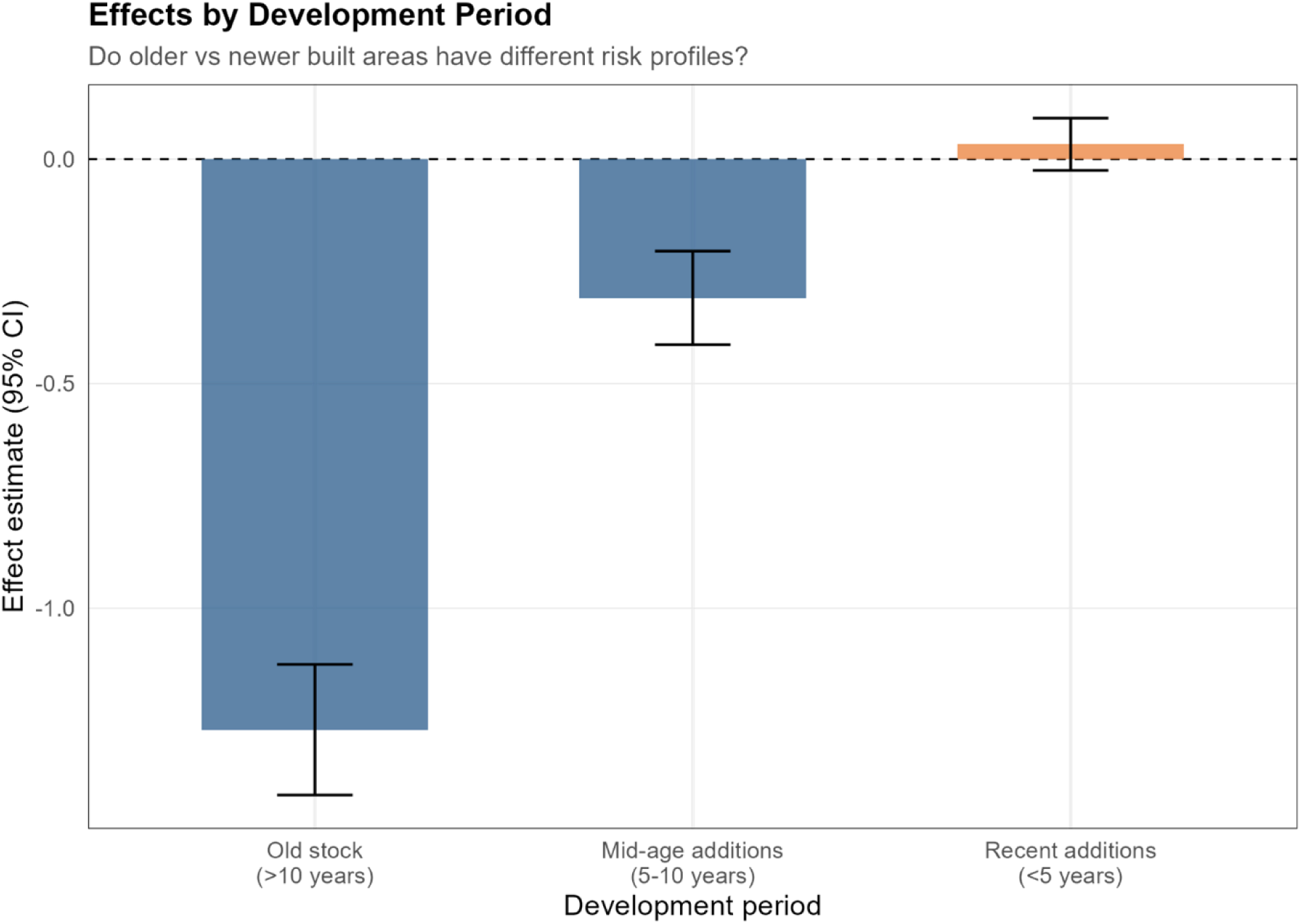
Lag model. Historical v recent urbanisation.

#### Distributed lag by development period

When built area was decomposed by development period, older urban fabric was protective, with effect sizes weakening as development became more recent. Built environment present >10 years ago had a strong negative association (β = −1.270, p < 0.001), mid-age additions (5–10 years ago) remained negative but smaller (β = −0.309, p < 0.001), and recent additions (<5 years) were near-null and not significant (β = 0.033, p = 0.267). This gradient is consistent with an infrastructure maturation mechanism in which protective benefits accrue over time rather than immediately following expansion (Figure 3).

**Figure 3.**
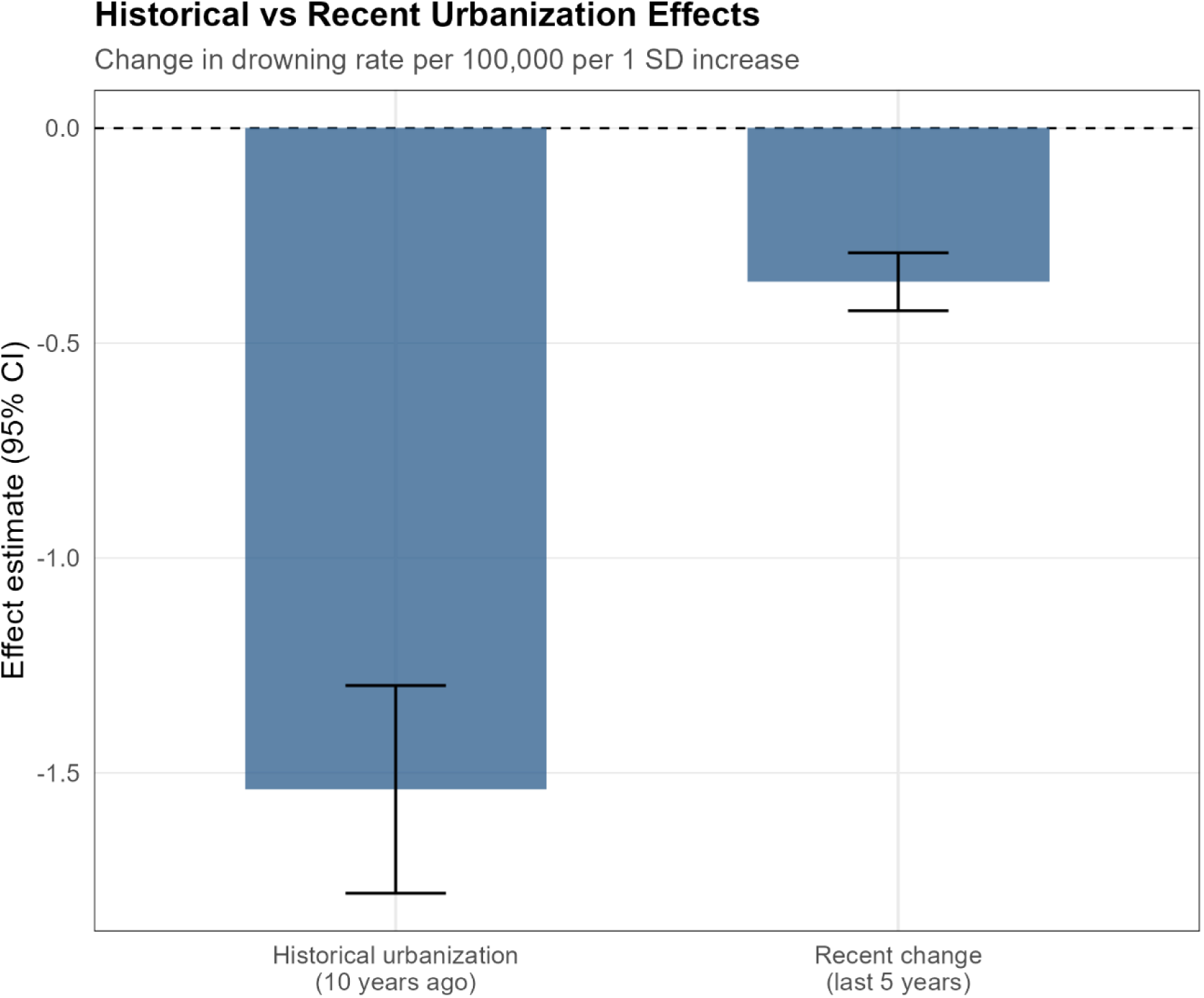
Lag model. Effects by development period.

#### Infrastructure timing effects

In the infrastructure timing model, a higher share of new development was associated with higher drowning mortality (β = 0.604, p < 0.001). Infrastructure lag was negative (β = −0.770, p < 0.001), and the new development × poor infrastructure interaction was negative but marginal and not statistically significant (β = −0.064, p = 0.055), providing limited evidence that the combination compounds risk. Crowding was not significant (β = 0.135, p = 0.202), while built area near water remained negatively associated (β = −0.616, p < 0.001).

#### Income-stratified temporal effects

Temporal associations differed markedly by income group (Table S8). In low-income settings, the crowding × growth interaction was strongly positive (β = 2.02, p < 0.001), while both the 5-year built change ratio (β = −1.84, p < 0.001) and recent development ratio (β = −1.09, p < 0.001) were negative. In lower-middle income countries, recent development ratio remained negative (β = −1.07, p < 0.05) and growth acceleration was more strongly negative (β = −0.38, p < 0.001), with a small negative association for growth volatility (β = −0.06, p < 0.05). In upper-middle income settings, patterns reversed: recent development ratio was strongly positive (β = 2.38, p < 0.001), built change ratio was strongly negative (β = −2.65, p < 0.001), growth acceleration was positive (β = 0.31, p < 0.001), and growth volatility was negative (β = −0.11, p < 0.001). In high-income countries, the crowding × growth interaction was negative (β = −1.13, p < 0.01) and recent development ratio was weakly positive (β = 0.32, p < 0.05), while growth volatility was negative (β = −0.12, p < 0.001). Overall, these results suggest that densification and new development may elevate risk in lower-resource contexts but can be neutral or protective where infrastructure and regulation are stronger, with upper-middle income countries showing the clearest indication that age of development was associated with higher risk (Figure 4).

**Figure 4.**
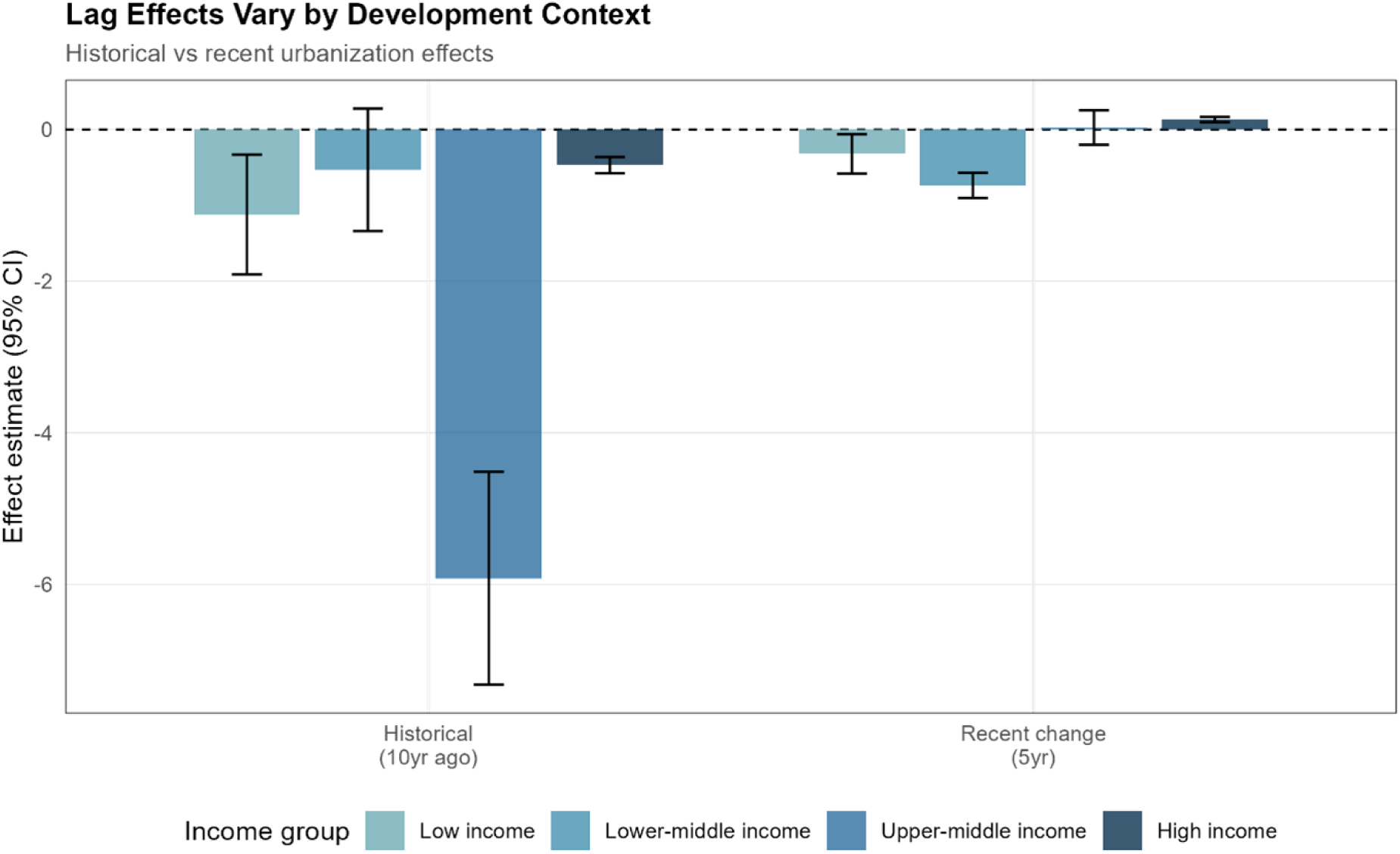
Lag effects by income group.

Income-stratified lag models also diverged (Table S8). Historical urbanisation level was negative in most strata, strongest in upper-middle income countries (β = −5.92, p < 0.001), and non-significant in lower-middle income countries (β = −0.53, n.s.). Recent urbanisation change was negative in low-income (β = −0.32, p < 0.05) and lower-middle income settings (β = −0.74, p < 0.001), near-null in upper-middle income countries (β = 0.03, n.s.), and positive in high-income settings (β = 0.13, p < 0.001), indicating that recent growth may be associated with higher risk only in the highest-income contexts (Figure 5).

**Figure 5.**
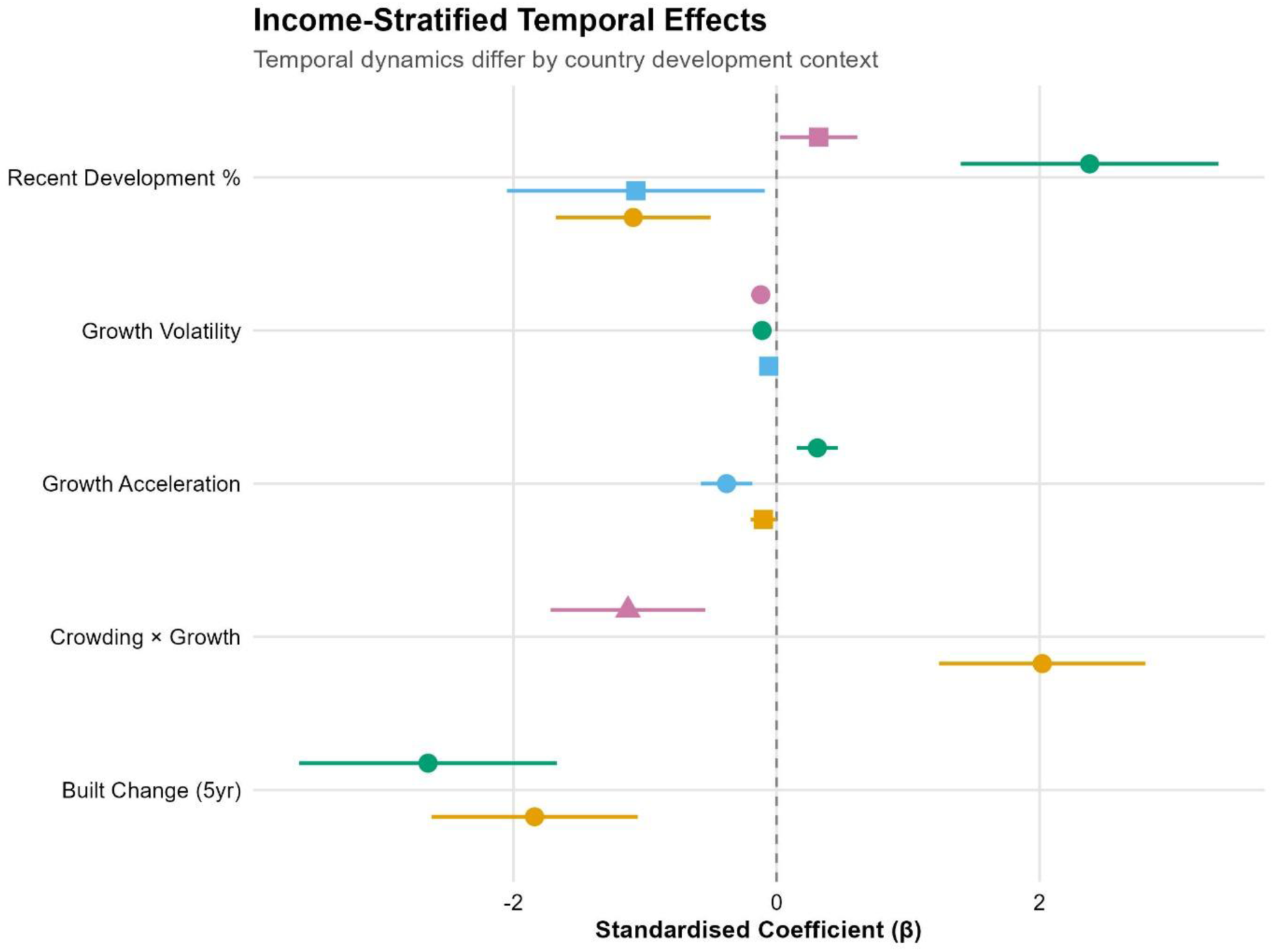
Income stratified temporal effects.

### Model diagnostics

Diagnostics indicated the main model performed well on core assumptions: multicollinearity was negligible (all VIFs <2) and the high ICC (∼0.98) supported a random-intercepts structure, with most variation occurring between regions rather than within regions over time (Tables S9). Model fit was weaker for distributional assumptions and effect heterogeneity: transformed outcomes fit better than the untransformed Gaussian, and random-slopes models improved fit but were not reliably estimable. Sensitivity checks were stable to sample restrictions (≥10 years) but somewhat influenced by extreme outcome values (winsorisation improved fit), suggesting results are broadly robust but primarily reflect between-region differences, with departures in extreme settings. Moran’s I was non-significant across all neighbourhood specifications (k = 3: I = 0.115, p = 0.109; k = 5: I = 0.057, p = 0.076; k = 8: I = −0.077, p = 0.392), and LISA identified no significant spatial clusters (100% of countries non-significant; Tables S9). These results confirm that region-level random intercepts adequately capture between-area variation, and spatially structured random effects are not required.

## Discussion

Our findings identify broad global associations between urban development history and drowning burden, suggesting that regions with older built environments tend to experience lower drowning mortality. These macro-structural associations should be interpreted alongside fine-scale spatial studies demonstrating that drowning risk frequently clusters around specific local water features and socio-demographic deprivation, which global models may smooth over. For example, spatial hotspot analyses in the UK [17] highlight localized clustering, while studies in Iran [18] emphasise the role of particular environmental water bodies, and work from British Columbia [19] identifies education and housing typologies such as apartments as significant predictors. Unlike hotspot detection approaches designed to identify geographically concentrated risk, our macro-structural modelling framework examines cross-national variation in development trajectories and infrastructure history. Importantly, these findings do not contradict prior rural–urban comparisons showing higher drowning rates in rural settings. Greater urbanisation was found to be generally protective. Rather, these findings indicate that within regions experiencing urban growth, newly developed or rapidly changing built environments may present distinct risks, even where long-established rural areas continue to have high drowning rates.

The observed associations between built environment age and drowning burden are ecological and should not be interpreted as evidence of a causal pathway. The high intraclass correlation coefficient indicates that most variation occurs between regions rather than within regions over time, suggesting that the identifying variation is driven primarily by cross-sectional differences rather than longitudinal change. Accordingly, these findings are consistent with the possibility that longer-established infrastructure, regulatory systems, or safety norms may contribute to lower risk [20], but they do not establish such processes directly. Infrastructure lag and development history may also operate differently across income contexts. In high-income settings, longer development histories may coincide with progressively strengthened building codes, water safety regulation, and service provision, whereas in lower-middle-income contexts, rapid urban expansion may outpace infrastructure and regulatory capacity, generating distinct risk profiles. These contextual differences further underscore that global models can obscure important socio-environmental heterogeneity.

While our focus was drowning, the broader relationship between urban form and injury risk has parallels in other domains. For instance, global analyses of urban design and road transport injury [21] demonstrate that structural features of the built environment are associated with injury burden across mechanisms, suggesting that urban development patterns may shape multiple injury risks simultaneously.

The policy implications of these findings relate to aligning urban development with water safety planning. In rapidly urbanizing lower– and middle-income settings, preventive strategies may include integrating water-safety considerations into land-use planning, enforcing barriers around high-risk water bodies, strengthening building codes related to drainage and flood mitigation, expanding community-based swimming and rescue training, and improving surveillance systems to detect emerging spatial clusters. In higher-income contexts, continued regulatory maintenance, retrofitting of aging infrastructure, and targeted interventions in socioeconomically deprived or environmentally exposed subregions may be warranted. By situating drowning prevention within broader infrastructure and development policy, our results support intersectoral approaches linking urban planning, housing, education, and injury prevention.

The distinction between high-income and low-/middle-income contexts is central to interpreting these findings, as development history, infrastructure capacity, and regulatory environments likely modify the association between built environment age and drowning risk, something that is consistent with previous literature int this space [22]. Future research integrating macro-structural modelling with subnational spatial analyses would further clarify how global development trajectories interact with local environmental exposures to shape drowning risk.

Limitations include the ecological design (i.e. these results give little insight into individual differences when it comes to drowning), residual confounding (i.e. we did not account for a range of factors, like policy changes for example that may influence drowning rates), and indirect measurement of infrastructure and hazards. Region-level random intercepts absorb time-invariant contextual factors including broad socio-economic conditions and national policy environments, but cannot isolate the effects of specific policy changes or sub-national socio-economic shifts occurring during the study period. Furthermore, while we have utilised the most granular GBD data available when it comes to drowning, the regions analysed here may still have had substantial heterogeneity. The observed effects may not hold where substantially smaller or larger areas are studied. Nighttime lights imperfectly capture service provision, as luminosity reflects electrification and economic activity rather than specific safety-relevant infrastructure. Bright urban cores may experience sensor saturation, while dim or informal settlements with limited electrification may be under-detected. As a result, lights may misclassify both highly serviced but low-light areas and densely populated but poorly serviced settlements, introducing measurement error that likely attenuates associations. For all datasets, resolution should be noted, particularly for GHSL data; with a 100m resolution this may have resulted is misclassified pixels in complex urban environments, for example. Similarly, the water proximity measure may underestimate exposure from seasonal, ephemeral, or small domestic water sources (wells, cisterns, ponds) that fall below satellite detection thresholds, and these unmapped exposures would attenuate rather than inflate observed associations. Furthermore, built-up measures at 5-year epochs may smooth short-term shocks and introduce measurement error. Smaller income-stratified samples also increase uncertainty within strata.

## Conclusions

Newly developing settings, especially those experiencing rapid change in dense contexts, may warrant incorporating water safety provisions into land-use planning, such as barriers around open water, drainage standards, and safe water access points, rather than assuming safety benefits accrue immediately with urban growth. Established high-density areas may require different approaches targeting crowding near water bodies. Given strong cross-context heterogeneity, priorities should be tailored to development context, though these recommendations should be interpreted cautiously given the ecological design, which identifies area-level associations rather than individual-level causal pathways. Future work should test non-linear specifications, incorporate richer measures of flooding/drainage and water access, and link remotely sensed change to documented infrastructure and policy timing to clarify mechanisms.

## Data Availability

All data used in this study is publicly available

